# CoVID-19 prediction for India from the existing data and SIR(D) model study

**DOI:** 10.1101/2020.05.05.20085902

**Authors:** Aditya Rajesh, Haridas Pai, Victor Roy, Subhasis Samanta, Sabyasachi Ghosh

## Abstract

CoVID-19 is spreading throughout the world at an alarming rate. So far it has spread over 200 countries in the whole world. Mathematical modeling of an epidemic like CoVID-19 is always useful for strategic decision making, especially it is very useful to gain some understanding of the future of the epidemic in densely populous countries like India. We use a simple yet effective mathematical model SIR(D) to predict the future of the epidemic in India by using the existing data. We also estimate the effect of lock-down/social isolation via a time-dependent coefficient of the model. The model study with realistic parameters set shows that the epidemic will be at its peak around the end of June or the first week of July with almost 10^8^ Indians most likely being infected if the lock-down relaxed after May 3, 2020. However, the total number of infected population will become one-third of what predicted here if we consider that people only in the red zones (approximately one-third of India’s population) are susceptible to the infection. Even in a very optimistic scenario we expect that at least the infected numbers of people will be 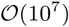.

## 1 Introduction

The outbreak of the novel coronavirus, CoVID-19, originated in Wuhan, China has spread rapidly across the globe. The spread of this infection is found to be so alarming that the World Health Organization (WHO) declared it as a pandemic disease on 11th March 2020 [1, 2]. This pandemic disease becoming a great threat to the health and safety of people across the globe due to its rapid spreading power and potential mortality. Presently, its evaluation is closely monitored by governments, researchers, and the public alike. Every country has limited medical resources, no matter how advanced the healthcare system. Moreover, in the absence of a vaccine, social distancing has emerged as the most widely adopted strategy to control the outbreak of the novel corona virus [3]. The Government of India has also announced a nation-wide lock-down starting from 24th March 2020 to reduce the contact rates in the population and slowing down the transmission of the virus. In recent times, epidemiological study for CoVID-19 data of India is started by a many research groups [4, 5, 6, 7, 8, 9, 10, 11, 12, 13, 14, 15, 16, 17, 18]. In this regard, classical epidemiological modeling framework [19, 20, 21], could be capable of the describing dynamics of CoVID-19 like a pandemic, and predicting the impact of mitigation measures currently undertaken by the Government of India where population density is very high.

One naive way to predict the immediate future of the progression of an epidemic is to fit the existing data with reasonable mathematical functions with a few free parameters. This has been done with the CoVID-19 India data and shown in Fig. (1). We choose the interval of March 12- April 3, 2020, to fit the total number of infected people with the following three parameters functions,

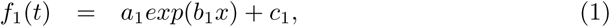

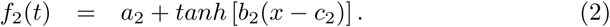

**Figure 1:**
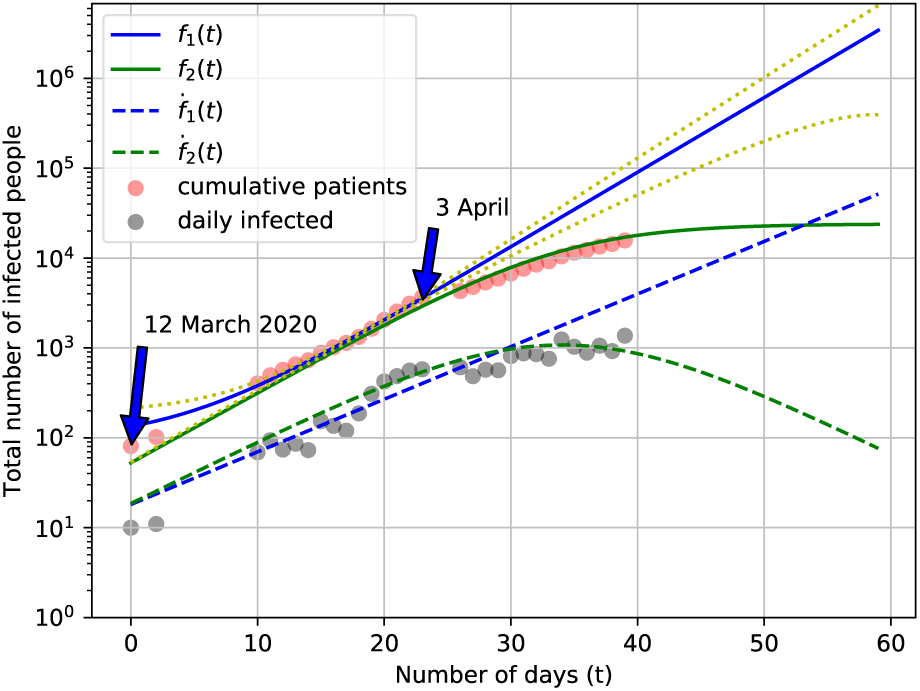
Functional fit to the measured data for the total infected people as a function of time (days). The best fit for *f*_1_(*t*) and its derivative was obtained by using python libraries. Two yellow dotted lines show the two sigma deviation from the best fitted values. The fit for *f*_2_(*t*) is done by eye estimation.

**Figure 2:**
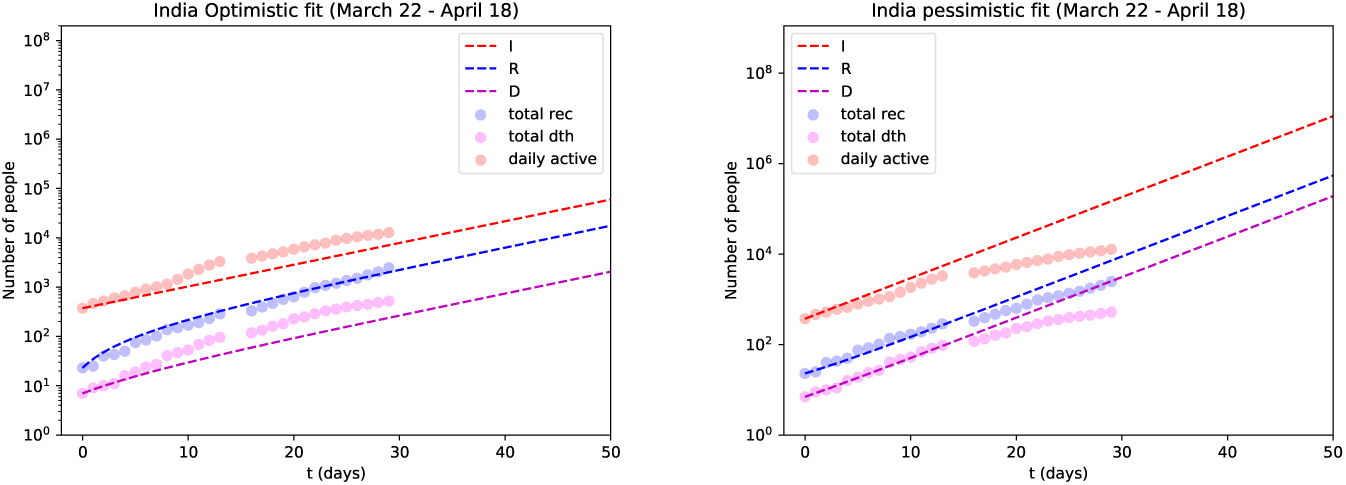
Comparison of SIR(D) model simulation (various lines) to the measured data (circles) for India during the period 22 March - April 18. The left panel corresponds the optimistic fit with *α* = 0.135, *β* = 0.03, and *γ* = 0.0035 and the right panel corresponds the pessimistic fit with *α* = 0.22, *β* = 0.01, and *γ* = 0.0035.

The motivation behind using an exponential function is well supported by the fact that for highly infection diseases like CoVID-19 on average a patient infects more than two people which leads to exponential growth. However, this rapid growth can be reduced by imposing conditions like lock-down or administering vaccines to the mass population, etc. and in that case, the initial exponential growth reduces and eventually settle to a constant value where no more new infections reported. This behavior of initial exponential rise saturating to a constant value is parameterized in Eq.(2). However, note that in this case Eq.(2) is not an unique function which shows this kind of behaviour, we can use other functional form such as 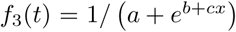 to represent the same behavior. Solid the blue and green line in Fig. (1) corresponds to the best-fit curve with the following values of the parameters *a*_1_ = 34 ± 8, *b*_1_ = 0.202 ± 0.010, c_1_ = (1.2 ± 0.5) x 10^2^ and *a*_2_ = 1.2 x 10^4^,b_2_ = 0.09, c_2_ = 34. As we can see both *f*_1_(*t*) and *f*_2_(*t*) seems to match the data well in the given period, then the question arises which case shall we believe? To investigate further, we study the derivative of these functions (shown by the dotted lines in Fig. (1)) which should give us the daily infected patients. We can again see that it is hard to distinguish the two scenarios from the data. However, after April 3 the rate of infected people reduces which also reflects in the derivative and here *f*_2_(*t*) better explain both data. Most likely this reduction in infection is due to the ongoing lock-down from March 24, 2020, the effect of which is seen after approximately two weeks as expected. But this kind of fitting the data and predicting the future has it’s own limitations, for example, the effect of future lock-down, vaccination, weather conditions that weakens the virus and reduce the virulent of the infection are hard to implement in this case. Also, the fact that recovered patients may act as herd immunity to stop the further spread of the infection is not included in this case. There are a few mathematical models available that incorporate some of the essential features and dynamics of an epidemic and can be used to study and predict the future of an epidemic like CoVID19.

In the present work, we use one of the effective mathematical model SIR(D) [22] that is simple yet quite effective to predict the future of the epidemic and the effect of a lock-down/ social isolation via a time-dependent coefficient of the model for India by using the existing data [23]. The data source used is from the website https://covid19india.org, which itself is sourced from WorldoMeters and other reliable sources.

## 2 Model description

The SIR(D) dynamical model is composed of four coupled differential equations to describe the progression of epidemiology. The model is built on the fact that on any given day the total population can be divided into mainly four different categories namely those who are susceptible to the infection (S), already infected people or active number of patient (I), number of recovered people (R), and the number of deaths (D). It is customary to neglect the usual daily birth and death rate which is also known as demographic. The SIR(D) model also assumes that the quantities S, I, R, and D are dependent only on time and not on the location. The time rate of change of these quantities are given by the following coupled ordinary differential equations

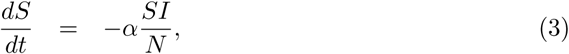

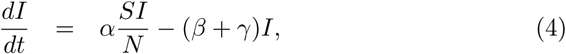

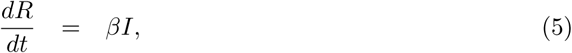

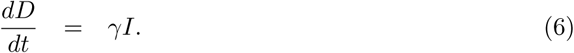

Here *α, β, γ* are the rates of infection, recovery, and mortality, respectively. The parameters *α* and *β* approximately represent the average number of contacts a person has during a single day and the reciprocal of the period for which an infected person remains infectious. It is important to check that the solutions of Eqs.(3-6) should satisfy the constraint *S*(*t*) + *I*(*t*) + *R*(*t*) + *D*(*t*) = *N*, where N is the total population. This is clearly seen as adding all four equations gives 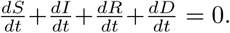. It is worthwhile to mention that for many infections, there is a significant incubation period during which individuals have been infected but are not yet infectious themselves. In this case one must take into account this fact while writing the differential equations, however, here we only discuss the SIR(D) model results. In this model, the ratio 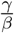 dictates the number of deaths to the number of infected people and hence it is named as fatality ratio [7].

Now according to the data from medical research all over the world, the CoVID-19 virus has a typical contagious period of around 8-10 days after infecting a person. This isn’t an exact figure and can be wrong by a few days give-or-take. This, however, gives us the ballpark figure for *α*. We can safely say that *α* should lie somewhere around ~ 0.1. The exact value can be obtained from the data at hand. Now for *β*, obtaining a ballpark may not be as simple. Since there are lots of factors that can affect how many people a single person meets in a day, like *α* it would be best to play with some values and see which one fits our data best. In this analysis, the value of *α* is fixed by matching the data.

## 3 Implementation

The data for this analysis is obtained from the CoVID19 India tracking website as mentioned earlier. The differential equations of the model were solved using the ODEINT functionality of the scipy module in Python. We also use the scipy module stat to calculate the Pearson correlation and p values.

## 4 Results

### 4.1 Prediction for India (ignoring lock-down)

Since there is no error reported in the data we could not carry out *χ*^2^ analysis at the moment and rely on the eye estimation for the best fit. But we should not try to find the best fit with the eye estimation alone as it is not a scientific way to obtain the best fit, rather we try to simulate two scenarios (1) optimistic where the fit underestimate the number of infected people and number of death but overestimate the number of recovered people (2) pessimistic scenario which is just opposite to the optimistic scenario. After tweaking the values of *α, β*, and *γ* in the SIR(D) model we find the current data for India is described well for the optimistic and pessimistic scenarios with the following values of the parameters

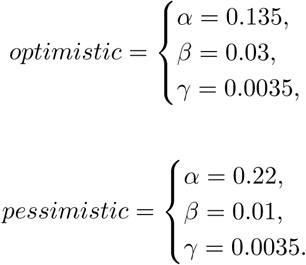

The left panel of Fig. (3) shows the comparison of the results obtained from the SIR(D) model for the optimistic case to the measured data for the duration of March 22 to April 17, 2020. Whereas, the right panel of Fig. (3) shows the same but for the pessimistic case. It is understood that these sets of parameters are not unique a different values of the parameter set *α*, *β*, and *γ* may also work well, but we checked that for a reasonable fit we can’t deviate too much from the values given above.

**Figure 3:**
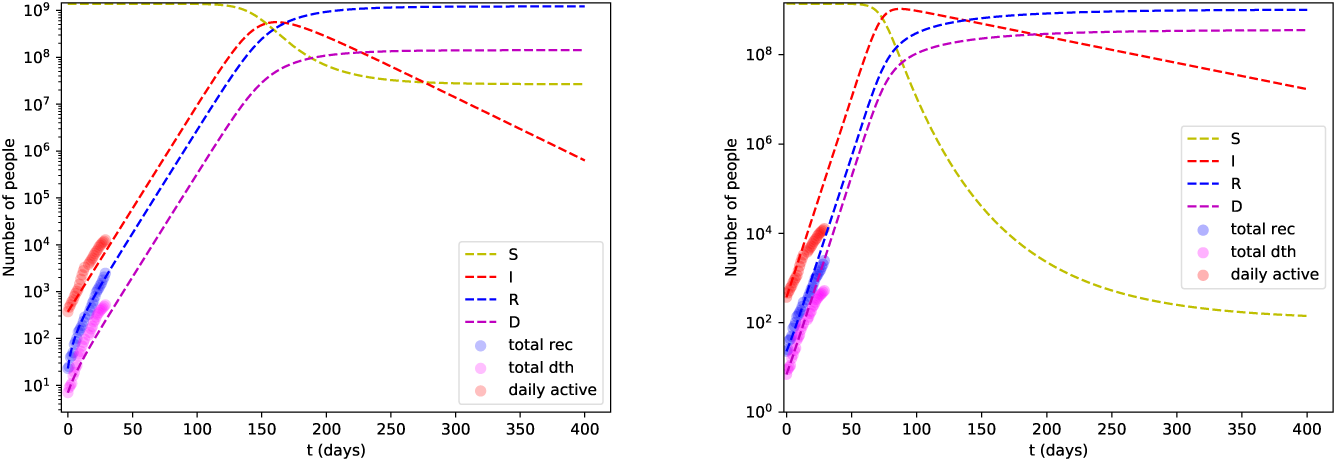
Prediction for India from the SIR(D) model simulation for two different scenarios and assuming no lock-down imposed. The left plot corresponds to the optimistic case wheras the right panel corresponds to the pessimistic case (see text for details).

As clearly seen, the data and model agree pretty well (except that the data shows sudden change in slope possibly originating due to the lock-down which we discuss later in details). To check the correlation, the Pearson correlation coefficient, and corresponding p-value were calculated (using the scipy package). The correlation coefficient for optimistic and pessimistic scenarios came out to be 0.99, 0.99,0.99 and 0.92,0.98,0.93 respectively (both cases indicate high positive correlation) for infected, recovered and death statistics, and the p-value came out to practically 0 (very small 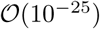, and 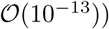 for infected and for recovered and the deaths 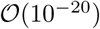, further indicating that the model captures the data trend very well. Going forward with these values of *α, β*, and *γ*, the plots for the extended period obtained from the model for the two scenarios are shown in Fig. (3). This model indicates that for optimistic case the peak of infection should come at around ~ 150 days from March 22, with the maximum number of infected people being close to ~ 200 million and approximately ~ 100 millions death and for the pessimistic case almost the whole population got infected (as can be seen from the suspected population) with the maximum number of people infected at around ~ 100 days from March 22.

However, we shall not take this result very seriously for various reasons, some of which are as follows:

1. although the data is taken during the lock-down period the actual effect of the lock-down is seen later (mostly from April 9), this important contribution of lock-down will be discussed in a later section,
2. some preliminary studies have shown that the virus becomes weaker in hot and humid weather, so the upcoming summer in India and the northern hemisphere will prohibit the spread to a certain extent,
3. some Indian states are doing very well in controlling the spread of the virus and hence the effective susceptible population for India is less than what is taken here.

We will try to elaborate on some of these aspects in the next section. It is also worthwhile to mention here that different states may reach peak infection at different times and hence the curve of infected people may be much flatter than the result obtained here provided the lock-down continues for some more times.

### 4.2 State-wise analysis

As mentioned earlier, the overall number of infected peoples, the peak height of the infection and it’s duration will largely depend on the performance of different states in controlling the epidemic. The reason is obvious since in the lock-down scenario the inter-state travel is banned and hence we can consider each state as a separate container where the infection progress at different rates. In an idealized situation where the lock-down continues unless all the states become infection free the total number of infected people in India will be much much smaller than predicted in the previous section because many states like Kerala, Goa, Odisha, Arunachal Pradesh, Sikkim, Manipur, Tripura, etc. are handling the situation quite well (as on 28 April 2020) and are expected to become virus free within next few months if lock-down continues. However, lock-down will likely be relaxed soon and people will travel from one state to another. Now, let us take two hardest-hit states of India Maharashtra and Madhya Pradesh. We try to explain the data for these two states from the SIR(D) model. the results are shown in Fig. (4). From the two cases, we can see that in both the states the infection will peak around 150 days from March 22, 2020.

**Figure 4:**
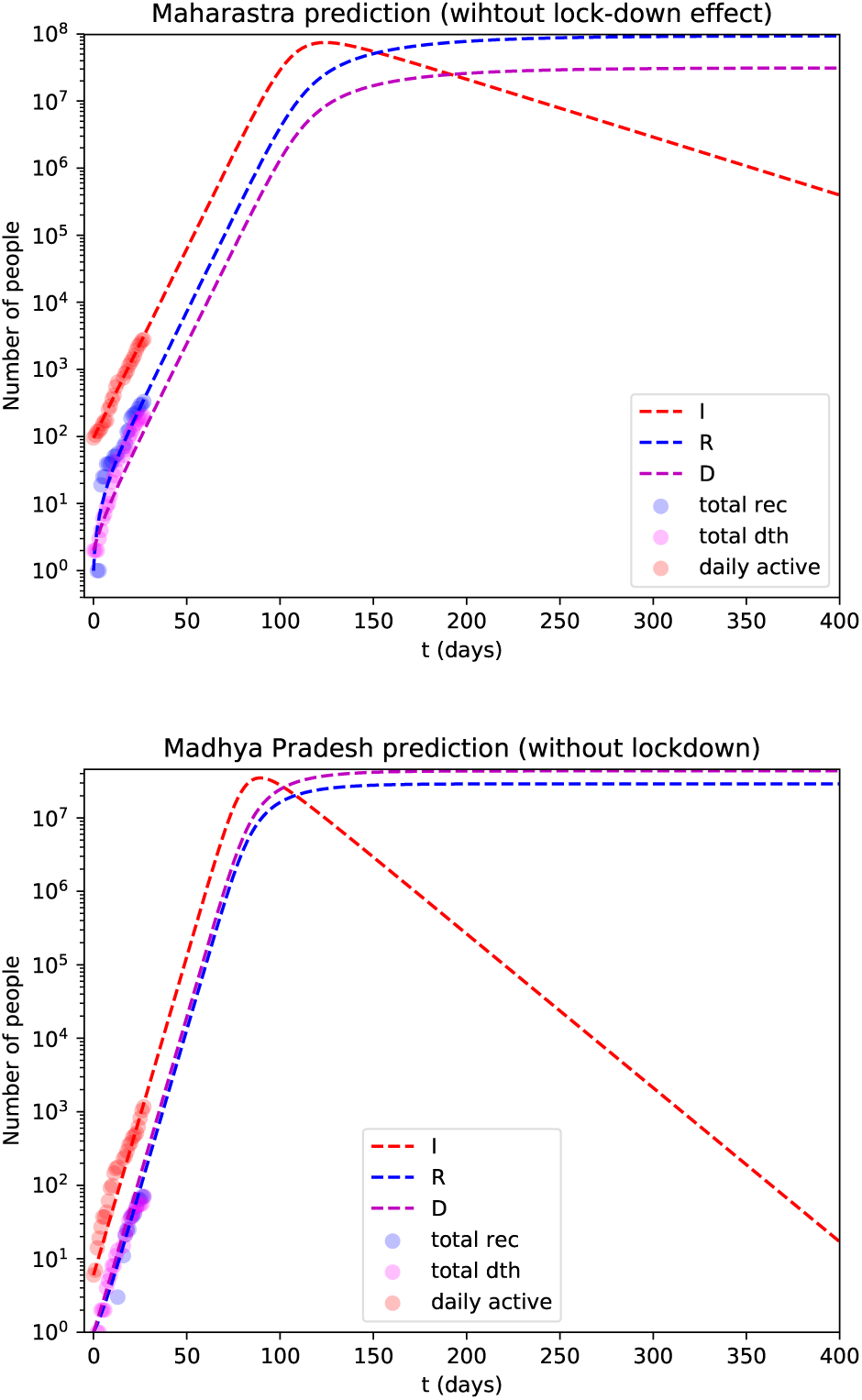
Predictions from the SIR(D) model simulation (mostly pessimistic prediction) for Maharashtra (top panel) and Madhya Pradesh (bottom panel). The values of the constants taken are *α* = 0.15, *β* = 0.015 *γ* = 0.005 for Maharashtra and *α* = 0.25, *β* = 0.02 *γ* = 0.03 for Madhya Pradesh.

In the above simulations for entire country or different states, we consider the whole population of the country/state to be susceptible to the infection. We can surely improve on that by subtracting the population of the green zones (where no active cases are found) from the total population to get the correct number of susceptible population. However, this is the most non-trivial task as there are many asymptomatic cases and it is hard to sharply distinguish between red and green zones. Hence, our assumption that the total population of a state or the country is susceptible to the infection may not be correct. Accordingly, for a lower number of susceptible population the number of infected people (and its peak value) will get reduced. In this context exploring the hot-spot zones with large outbreak, as defined by Government authority, might be an important input for further investigations. As an example, the hot-spot districts of Maharashtra (MH) with large outbreak and their corresponding populations [24] are listed in Table 1. If we consider only these hot-spot zones as a total box size for MH, then 50% population size will be reduced. From the hot-spot district to more microscopic zones might provide us more reduced box sizes, for which we can get a reduced peak value of infection for MH. Although micro-tuning this box size is real challenging task, which is not attempted in the present article, but planned for a future investigations.

**Table 1:** MH: Population of different red-zone (with large outbreak) disctricts in unit 10^6^

|  |  |  |  |  |
| --- | --- | --- | --- | --- |
|  | Mumbai | Pune | Thane | Nagpur |
| *Total Population* | 3.3 | 7.2 | 8.1 | 4 |
|  | Yavatmal | Aurangabad | Buldhana | Mumbai Suburban |
| *Total Population* | 2 | 2.8 | 2.2 | 8.5 |
|  | Sangli | Ahmednagar | Nasik | Total (MH) |
| *Total Population* | 2.5 | 4 | 4.9 | 49.5 |

### 4.3 Effect of lock-down and social distancing

To see the effect of lock-down in this model we assume that the rate of infection gets reduced due to the lock-down over the whole lock-down period. Since there are numerous effects such as local variation of the success of lock-down, the population density difference between urban and rural areas, etc., it is really hard to get an estimate about the real variation of the parameter *α* which primarily governs the rate of infection. The values of *β* and *γ* are mostly kept constant unless stated otherwise as to the recovery rate and the death rate hardly depends on the lock-down.

**Figure 5:**
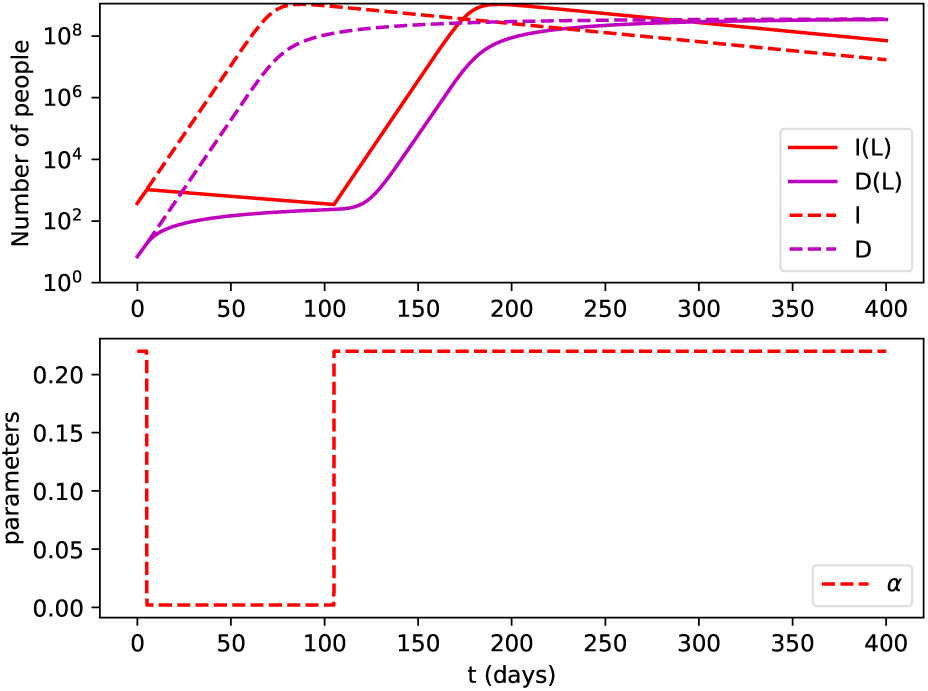
Effects of a 100 days ideal lock-down according to the SIR(D) model simulation. The top panel shows the I(t), R(t), and D(t) obtained for lock-down (solid lines) and without it (dashed lines). The bottom panel shows the time-dependent *α*, we arbitrarily reduce the value of *α* by a factor of 100 from its original value 0.22 (without lock-down).

In Fig. (5) we show the effect of an ideal 100 days long lock-down as per the SIR(D) model on the progression of the CoVID-19 in India. We call it ideal because we choose the values of α during the lock-down almost hundred times smaller than the observed value which means the number of infected people gets reduced during the lock-down period (as seen from the figure). This certainly is not the case for India where the data does not support this fact. Some of the media is showing this kind of effect of lock-down which we think is not applicable for highly infectious CoVID-19 disease.

**Figure 6:**
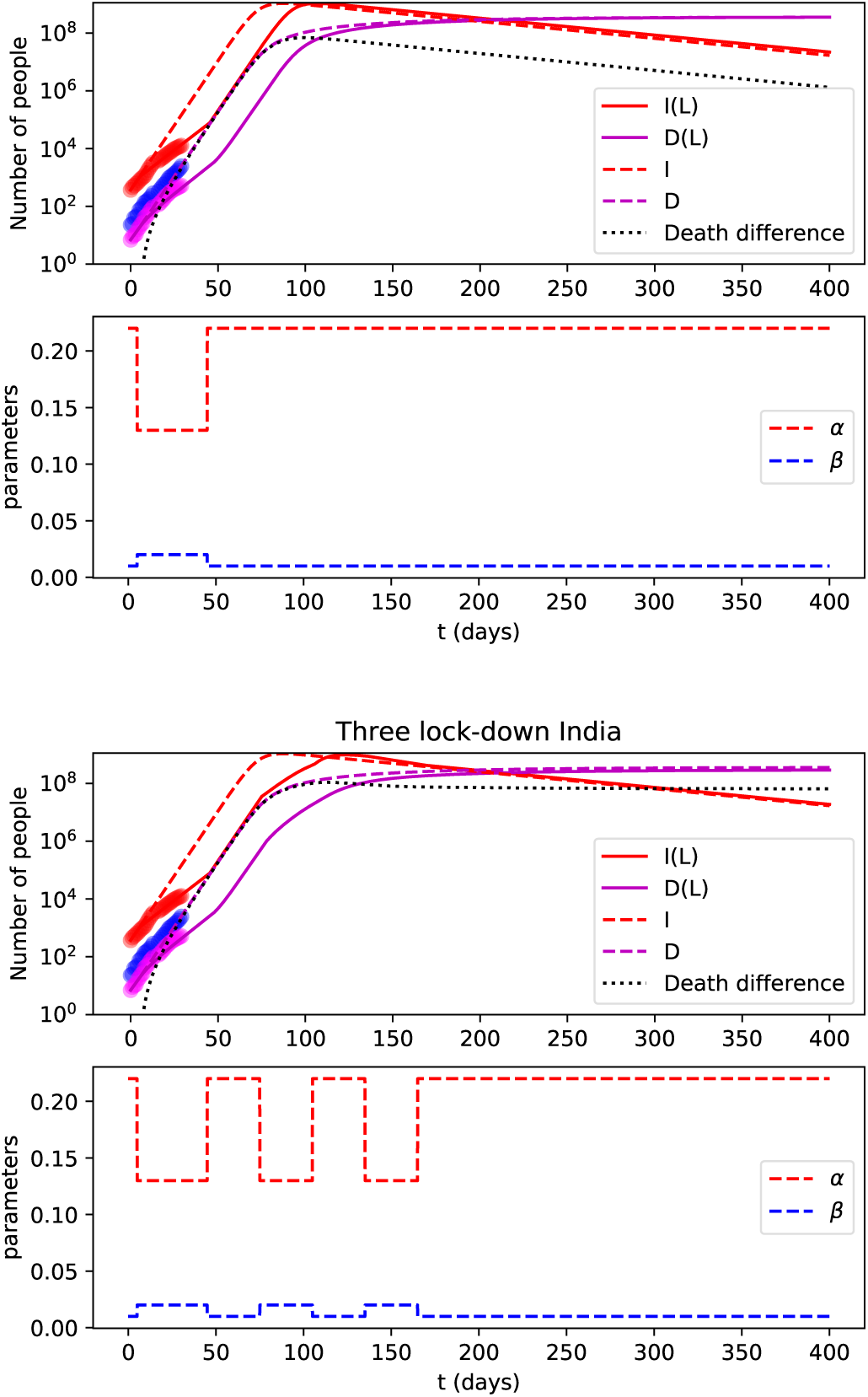
Top most panel shows the effect of a single 40 days long lock-down according to the SIR model simulation (different lines). The second top-most panel shows the temporal variation of *α* and *β* adjusted according to the lock-down duration to best match the observed data (shown by circles). The bottom two figures show the same thing but for three lock-down scenario (see text for details).

Results for a more realistic situation is shown in Fig. (6). The top panel of Fig. (6) shows the results for a single lock-down imposed for 40 days (same as ongoing lock-down in India) and here we choose a more reasonable time-dependent values of *α* and *β* which describes the actual data. Here it is clear that even imposing the lock-down does not make 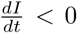 as was seen for the previous case of ideal lock-down. However, the lock-down surely delays the appearance of the peak by ~ 50 days or more, a crucial time required for the Government to be more prepared. Also, if a working vaccine arrive in the market this lock-down will definitely save millions more lives.

Lastly, we show effect of three lock-downs lasting for (40-30-30) days and between any two lock-down there is one month gap. The corresponding results are shown in the bottom panel of Fig. (6). According to these results there is hardly any noticeable changes to the peak position of infection between one lock-down scenario and a multiple lock-down scenario. However, we must be more careful when analysing data spanned between many orders of magnitudes in values. One important finding related to the above mentioned fact is that the difference in number of deaths (shown by the black-dot lines in Fig. (6) between these two scenarios is quite large, for example, during the peak of the infection, the difference in number of deaths can be as large as 10^6^ between the single lock-down and the multiple lock-down scenario.

## 5 Concluding Remarks

The present analysis of the SIR(D) model indicates a scarily high number of peak infections for India by comparing the existing data to the model simulation. However, what one must note, is that the parameters chosen for the model aren’t unique. For example, with measures like lock-downs and quarantines, the contact between people will reduce drastically, indicating that α will reduce to a lower value used here. Consequently, the peak would reduce significantly. According to the model study the peak will appear after approximately 100 days from March 22, 2020, i.e., around the last week of June 2020. We also show the difference between a single and multiple lock-down scenario and found that a large number of deaths may be prevented by imposing multiple lock-downs with interim one month no lock-down periods. Discovery of a working vaccine will of course drastically change the current trend, also hot and humid upcoming summer and monsoon season will provide some barrier to the virus spread. Our work also suggests that the announcement and implementation of a nation-wide lock-down by the Government of India was a right and courageous decision to contain COVID-19.

The present acute crisis demands that in future researchers from various disciplines need to collaborate and work together with doctors, nurses, politicians, and public administrators to fight against future pandemics. Last but not least, human civilization has witnessed and survived many devastating pandemics during the past few millenniums and this time it will also do the same. Now, we have advanced technology and medical facilities which will help us to fight against the new pandemic.

## Data Availability

All data are available in the given link below.

https://www.covid19india.org/

## 6 Acknowledgement

H. P. is grateful for the support of the Ramanujan Fellowship research grant under SERB-DST (SB/S2/RJN-031/2016), Government of India. V.R. acknowledges financial support from the DST through INSPIRE Faculty research grant, Government of India. VR also like to thank Prof. Sourendu Gupta for an initial discussion on SIR(D) model. SS acknowledges financial support from Polish National Agency for Academic Exchange. For group discussion through whats-App, Skype and Facebook, authors are thankful to other members of covid19-QGP group - Santosh K. Das, Balbeer Singh, Ashutosh Das, Akash Gupta, Souvik Paul, Ankit Anand, Ankita Mishra and Vivek.

